# Response to Tozinameran (BNT162b2) booster in twice-vaccinated kidney transplant and maintenance dialysis patients

**DOI:** 10.1101/2021.10.20.21264403

**Authors:** Iddo Z. Ben-Dov, Keren Tzukert, Michal Aharon, Hadas Pri-Chen, Yonatan Oster, Esther Oiknine-Djian, Dana G. Wolf, Michal Dranitzki Elhalel

**Affiliations:** Departments of Nephrology, Hadassah Medical Center, Faculty of Medicine, Hebrew University of Jerusalem, Israel; Clinical Microbiology and Infectious Diseases, Hadassah Medical Center, Faculty of Medicine, Hebrew University of Jerusalem, Israel

**Keywords:** COVID-19, viral infections, booster, vaccines, end stage renal disease, kidney transplantation, hemodialysis, peritoneal dialysis

## Abstract

**Background:** Patients needing chronic RRT are at risk for severe COVID-19 and mount a lesser response to mRNA vaccination. We describe the impact of booster administration in these patients, amidst a third wave of infections and deaths.

**Methods:** In the setting of a prospective COVID-19-centred cohort study in dialysis and kidney transplant patients we examined humoral responses to booster vaccination, and subsequent infection risk.

**Results:** We quantified antibodies (DiaSorin) in 198 maintenance dialysis patients, 314 kidney transplant patients and 82 controls, without prior COVID-19 infection. Prior to boosting, 79% of controls, 35% of dialysis and 11% of transplant patients had levels ≥59 AU/ml (putatively protective), while 8-54 days after a third injection, respective rates were 100%, 93% and 58%. Risk factors for antibodies <60 AU/ml despite booster injection were transplant vs dialysis, OR=18.4 (p<0.0001), transitioning from dialysis to transplantation or vice versa, OR=15.7 (p<0.01) and days post injection, OR=0.953 (p<0.05). Antibody step-up after the booster inversely correlated with the second dose step-up (r=-0.33, p<0.01). In this surge, 2 controls, 2 dialysis and 9 transplant patients had COVID-19. Antibody level ≥59 AU/ml at any time point independently associated with reduced risk of infection during this surge, OR=0.264 (p=0.048).

**Conclusions:** We show that a third dose of tozinameran boosts antibody levels in patients receiving RRT, as it did in controls. Most patients have now reached antibody levels likely to protect from infection. Antibodies were higher after the third dose compared to previous peaks, which may hint that the latest immune response may be more robust and sustainable, even in immune-compromised patients. Kidney transplant recipients showed the most striking enhancement, exhibiting >5.5-fold increase in the percentage of patients with protective antibody levels. However, many transplant patients remain below threshold even after boost injection, necessitating further boosting strategies.

## Introduction

On 22-Sep-2021 the U.S. Food and Drug Administration amended the emergency use authorization for the Pfizer-BioNTech COVID-19 Vaccine (tozinameran) to allow the use of a single booster dose, to be administered at least six months after completion of the primary series in certain populations including individuals 18 years of age and older at high risk of severe COVID-19^1^. However, boost vaccination of solid organ recipients commenced in Israel as early as mid-July. Additional at risk populations, including elderly individuals, patients treated with maintenance dialysis and healthcare workers trailed briefly; the general adult population soon followed, resulting in increased protection from infection^2^. Patients needing chronic renal replacement therapy are at risk for severe COVID-19^3^ and have been shown to mount a lesser humoral response to mRNA vaccination^4^. In our single center cohort we have observed that reduced vaccine-elicited antibody levels in dialysis and kidney transplant patients associated with increased infection rates^5^. Herein, we describe the short term impact of booster administration in these patients, amidst a third wave of infections and deaths currently experienced in Israel primarily by the Delta variant, possibly amplified by waning immunity (**Figure 1**) ^6^.

**Figure 1.**
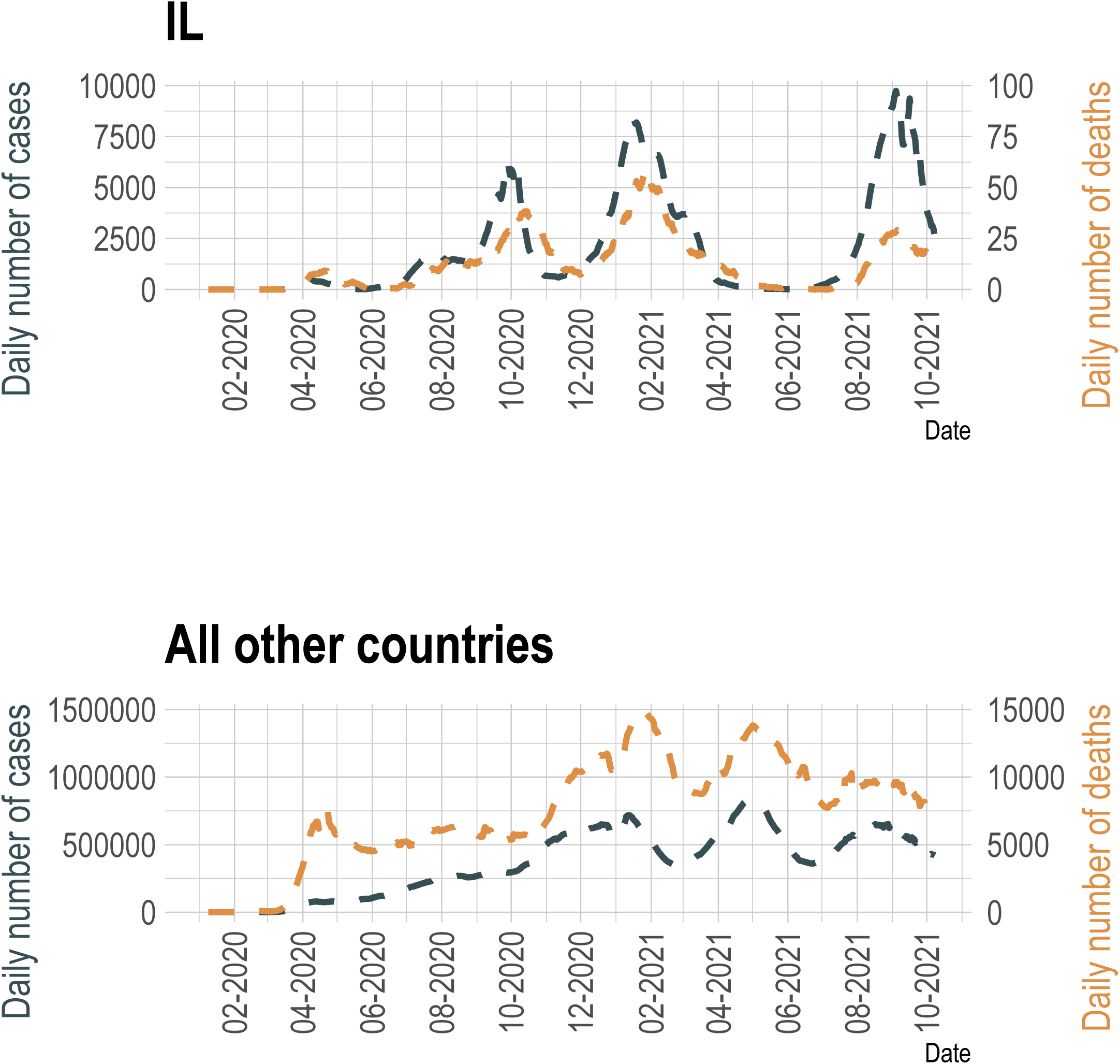
Daily number of COVID-19 cases (dark green) and deaths (light brown) in Israel (**top**) and rest of the world (**bottom**) according to data downloaded 09-Oct-2021 from the World Health Organization Coronavirus Dashboard^6^. Seven-day moving averages were plotted using the ggplot2 and tidyquant R/Bioconductor packages.

## Participants and Methods

Prior to initiation of vaccination we launched the COVID-19 mRNA Vaccine Immunogenicity in patients with end stage Renal Disease (COVIReD) prospective cohort study designed to investigate the long-term kinetics and implications of antibody response to COVID-19 vaccine and infection in this vulnerable population. Its methods and preliminary results have been recently published^5^. In this update, we report patients’ humoral responses to booster vaccination. Anti-SARS-2 antibodies were quantified using the LIAISON SARS-CoV-2 S1/S2 IgG test (DiaSorin, Sallugia, Italy). Risk factors for low antibodies were assessed using a generalized linear mixed effects model accounting for repeated measurements (lme4^7^ on the R statistical platform^8^). Infection risk was modelled with logistic regression, being that many events occurred over a short time interval.

## Results

In total, we quantified anti-S1/S2 IgG levels at multiple time points in 198 maintenance dialysis patients, 314 kidney transplants and 82 controls not previously infected with COVID-19, of whom 65, 87 and 15 respectively had been tested after receiving a vaccine booster. **Figure 2a** shows the individual and median values across study groups and vaccination episodes.

**Figure 2:**
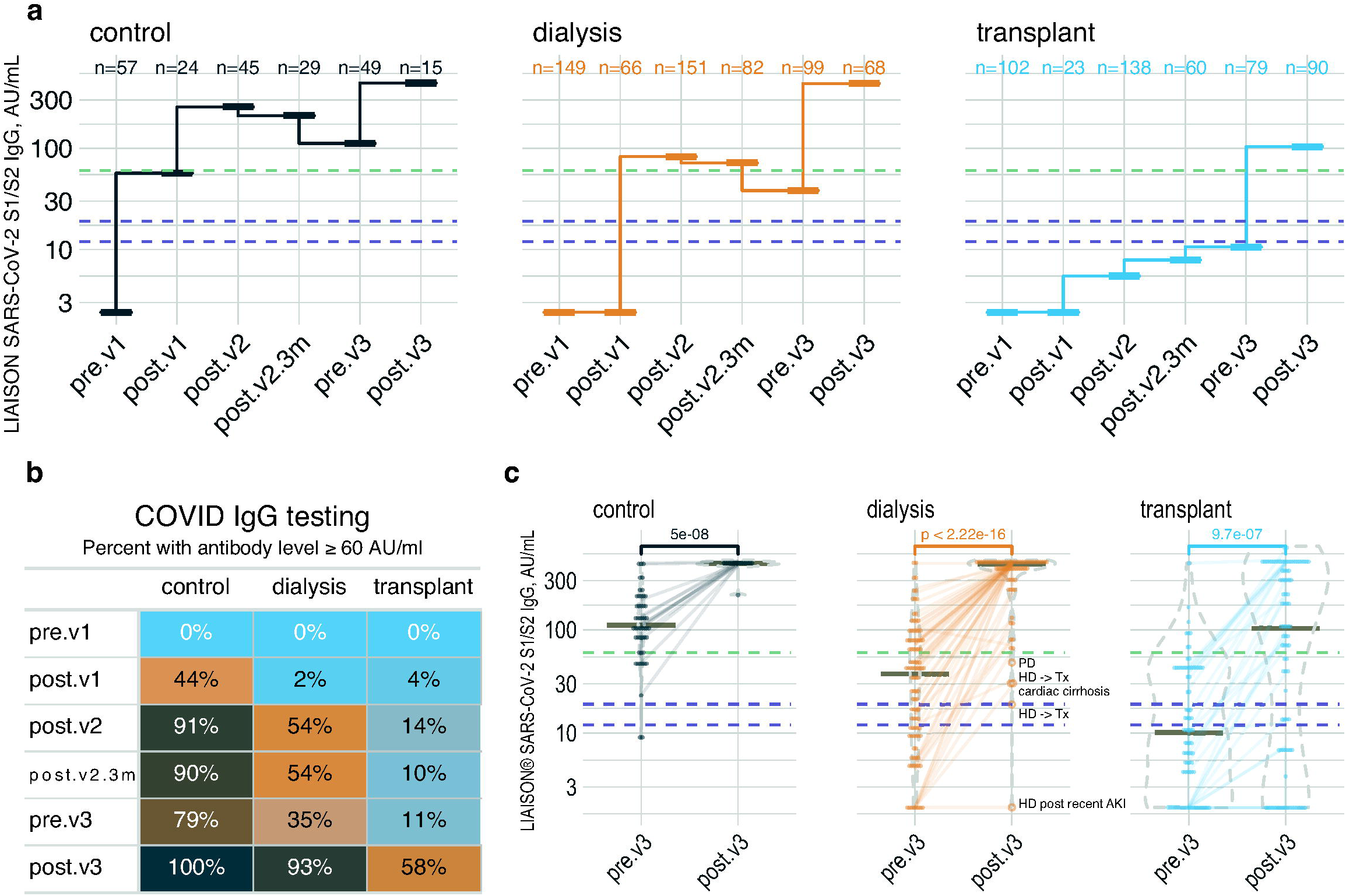
Antibody levels repeatedly measured across study groups and time points in patients not previously infected with COVID-19. (**a**) Median plots showing the summarized antibody level in a logarithmic scale in controls (left), dialysis patients (middle) and transplant patients (right). The dashed purple lines represent the equivocal range (12-19 AU/ml), while the dashed green line represents our proposed protective threshold, 59 AU/ml. (**b**) The percentage of participant with threshold or higher antibody level at each tome point, by group. (**c**) Dot and violin plots, with horizontal lines at medians, focused on the latest study time points. Semi-transparent lines connect repeated measurements from the same participant. Time point abbreviations: pre.v1, before vaccination; post.v1, after a single jab; post.v2, up to 10 weeks after the second dose; post.v2.3m, 3-5 months after double vaccination; pre.v3, 6 months after double vaccination; post.v3, 8-54 days after the booster shot.

We have previously noted that with a most recent antibody level at or above 59 AU/ml no post-vaccination cohort participant became infected with COVID-19, suggesting a protective cutoff^5^. **Figure 2b** shows that prior to boosting, this level was achieved by 80% of controls, 35% of dialysis patients and 10% of transplant patients, while 8-54 days after booster injection respective rates were 100%, 93% and 58%. Risk factors for antibody levels < 60 AU/ml despite booster injection (control subjects excluded) were being in the transplant group, OR=18.4 vs. dialysis (p<0.0001), transitioning *from* dialysis to transplantation during the study period or *vice versa*, OR=15.7 (p<0.01, **Figure 2c**) and time post injection, OR=0.953 per day (p<0.05). Age and sex were insignificant. Curiously, in a subset of 82 participants from all subgroups with available measurements before and after each vaccination, antibody step-up stimulated by the booster dose inversely correlated with the step-up after the second dose (r=-0.33, p<0.01). Finally, thirteen participants were diagnosed with COVID-19 since early July 2021; 2 controls, 2 dialysis patients and 9 transplant patients. Achieving an antibody level ≥60 AU/ml at any time point during the study associated with reduced risk of infection during the current surge, OR=0.264 (p=0.048), according to a logistic regression model also including age and the number of vaccinations.

## Discussion

Vaccination against COVID-19 was widely hoped to be the silver bullet against this deadly pandemic. However, starting July 2021, six months after mass vaccination in Israel, new infections surge, affecting also twice-vaccinated persons. In this report we show that a third dose of the BNT162b2 vaccine can effectively boost the antibody levels of high-risk chronic renal replacement therapy patients, as it did in healthcare controls. Most patients have now reached an antibody level likely to associate with protection from infection. Interestingly, the anti SARS-CoV-2 IgG levels were higher after the third dose than after the previous doses which may hint that the latest immune response may be more robust and hopefully sustainable, even in immune-compromised patients. In fact, kidney transplant recipients showed the most striking enhancement in humoral response to the booster dose, exhibiting an almost 6 fold increase in the number of patients developing a protective level of antibodies, greater than previously described^9^. However, many transplant patients remain below threshold in terms of antibody levels even after boosting. In our transplant clinic we did not lower immunosuppression prior to the booster dose, although various transplant centers contemplated transiently reducing immunosuppression to facilitate response (personal communications). Our experience is consistent with “real life” practice and may be of use to guide a practical recommendation for kidney patients receiving immunosuppression or dialysis.

## Conclusions

We show that a third dose of tozinameran boosts antibody levels in patients receiving RRT, as it did in controls. Most patients have now reached antibody levels likely to protect from infection. Antibodies were higher after the third dose compared to previous peaks, which may hint that the latest immune response may be more robust and sustainable, even in immune-compromised patients. Kidney transplant recipients showed the most striking enhancement, exhibiting >5.5-fold increase in the percentage of patients with protective antibody levels. However, many transplant patients remain below threshold even after boost injection, necessitating further boosting strategies.

## Data Availability

Data are available upon resaonable request

## Author Contributions

- Study conception and design – IZB-D, DGW, MDE
- Data acquisition including patient recruitment – MA, KT, HP-C, EOD
- Data analysis – IZB-D
- Data interpretation – IZB-D, KT, HP-C, YO, MDE
- Drafting the manuscript – IZB-D, KT, HP-C, YO, MDE
- Revising the manuscript – IZB-D, KT, HP-C, YO, EOD, DGW, MDE

## Acknowledgments

None

